# Usability of an At-Home Tablet-Based Cognitive Test in Older Adults With and Without Cognitive Impairment

**DOI:** 10.1101/2023.09.18.23295763

**Authors:** Thomas A. Bayer, Yanchen Liu, Igor Vishnepolskiy, Day Baez, Lisbeth Sanders, Rebecca Williams, Jeff Sanders, Anthony Serpico, Stefan Gravenstein

## Abstract

Mobile device-based cognitive screening has the potential to overcome the limitations in diagnostic precision and efficiency that characterize conventional pen and paper cognitive screening. Several mobile device-based cognitive testing platforms have demonstrated usability in carefully selected populations. However, the usability of take-home mobile device-based cognitive screening in typical adult primary care patients requires further investigation. This study set out to test the usability of a prototype mobile device-based cognitive screening test in older adult primary care patients across a range of cognitive performance. Participants completed the St. Louis University Mental Status Examination (SLUMS) and then used a study-supplied mobile device application at home for 5 days. The application presented 7 modules: 5 digital adaptations of conventional cognitive tests, 1 game-like experience, and 1 free verbal response module. Participants completed the System Usability Scale (SUS) after using the application. A total of 51 individuals participated, with a median (IQR) age of 81 (74–85) years. Cognitive impairment (SLUMS score < 27) was present in 30 (59%) of participants. The mean (95% Confidence Interval [CI]) SUS score was 76 (71–81), which indicates good usability. Usability scores were similar across ranges of cognitive impairment. SLUMS score predicted early withdrawal from the study with an area under the receiver operating characteristic curve (95% CI) of 0.78 (0.58-0.97). Take-home mobile device-based cognitive testing is a usable strategy in older adult primary care patients across a range of cognitive function, but less viable in persons with severe cognitive impairment. Take-home mobile device-based testing could be part of a flexible cognitive testing and follow-up strategy that also includes mobile device-based testing in healthcare settings and pen-and-paper cognitive testing, depending on patient preferences and abilities.

**AUTHOR SUMMARY:** Performance-based cognitive screeners play a critical role in the identification, triage, and management of persons with Major Neurocognitive Disorder in primary care, neurology, and geriatric psychiatry. Commonly used tests consume valuable medical provider time, can be unpleasant for patients, and provide minimal information about specific domains of cognition. Cognitive testing on a take-home mobile device could address these limitations. We tested the usability of a prototype cognitive testing application using take-home devices in 51 older adult primary care patients across a range of cognitive function. Participants found that the application had good usability, but more severe cognitive impairment predicted voluntary withdrawal from the study. These findings establish that take-home mobile device-based cognitive testing is usable among older adult primary care patients, especially those with less severe cognitive impairment.

## INTRODUCTION

Conventional cognitive screening methods have exhibited limitations in effectively capturing the intricacies of cognitive functioning among older adults. These methods often rely on in-person, pen-and-paper assessments, which can strain the already limited time available to healthcare providers in primary care settings.(1) Furthermore, these traditional methods may not fully capture the nuances of cognitive decline, especially in its early stages, potentially leading to missed opportunities for early intervention.(2) These screenings are often conducted in controlled clinical environments, which might not accurately reflect real-world cognitive performance in individuals’ day-to-day lives.(1) Such limitations can hinder the accurate detection and tracking of cognitive impairment, thereby underscoring the need for innovative approaches that harness the capabilities of modern technology to provide more nuanced and accessible assessments.

Screening for cognitive change sooner may lead to eligibility for new drugs, participation in clinical trials, deployment of meaningful interventions, and overall better health care outcomes. Early detection of major neurocognitive disorders enables more timely deployment of pharmacologic and non-pharmacologic interventions to help both persons living with dementia (PLWD) and their professional or family caregivers. Cognitive screening tools must also become more inclusive for demographically diverse individuals. A body of prior work has documented limitations of screenings that are not sensitive to varying socioeconomic, cultural, racial, or other differences.(3–9)

Although investigators have increasingly used mobile devices for cognitive testing in older adults, we lack evidence on at-home tablet-based cognitive testing in older adults requiring active participation.(10) A limited number of studies have demonstrated that various digital cognitive tests perform well in detecting dementia and mild cognitive impairment (MCI).(1) Although a study of tablet-based cognitive assessments found high usability ratings in older adults in a controlled setting, we need further study about the usability of such testing in a take-home format.(11) A study of a self-downloaded cognitive test demonstrated feasibility in users of an online citizen science platform. However, we must know the feasibility of at-home digital cognitive testing in an older adult primary care population.(12) We present the LifeBio Brain Phase 1 study. This study aimed to assess the usability of a take-home mobile tablet-based cognitive test in older adults who visit a geriatric primary care practice, with and without cognitive impairment.

## RESULTS

Of the 51 participants who provided informed consent, the median age was 81, with an intraquartile range (IQR) of 74 to 85. Self-reported race and ethnicity were White and not Hispanic or Latino in 51 (100%) of participants. Self-reported gender was female in 30 (59%) of participants. The Median (IQR) St. Louis University Mental Status Examination (SLUMS) was 25 (21–28).(13) Table 1 summarizes the characteristics of participants.

**Table 1:** Participant characteristics.

| **Characteristic** | **N=51** |
| --- | --- |
| Age, Median (IQR) | 81 (74-85) |
| 65-74, No. (%) | 13 (25) |
| 75-84, No. (%) | 25 (49) |
| 85-95, No. (%) | 13 (25) |
| Race: White, No. (%) | 51 (100) |
| Not Hispanic/Latino, No. (%) | 51 (100) |
| Female, No. (%) | 30 (59) |
| SLUMS Score, Median (IQR) | 25 (21-28) |
| 27-30, No. (%) | 20 (39) |
| 21-26, No. (%) | 20 (39) |
| 8-20, No. (%) | 10 (20) |
| Missing, No. (%) | 1 (2) |

Of the 51 individuals who consented to participate, 9 (18%) voluntarily discontinued before completing the study. One of these participants stopped participation before completing the SLUMS.

The mean (95% confidence interval) System Usability Scale (SUS) rating was 76 (71–81) overall. The mean SUS ratings were similar across the SLUMS score categories (Table 2). Validation of the SUS has determined that scores in the range of 73 to 85 represent ‘good’ usability.(14) The Pearson correlation coefficient for SUS and SLUMS was -0.03.

**Table 2:**
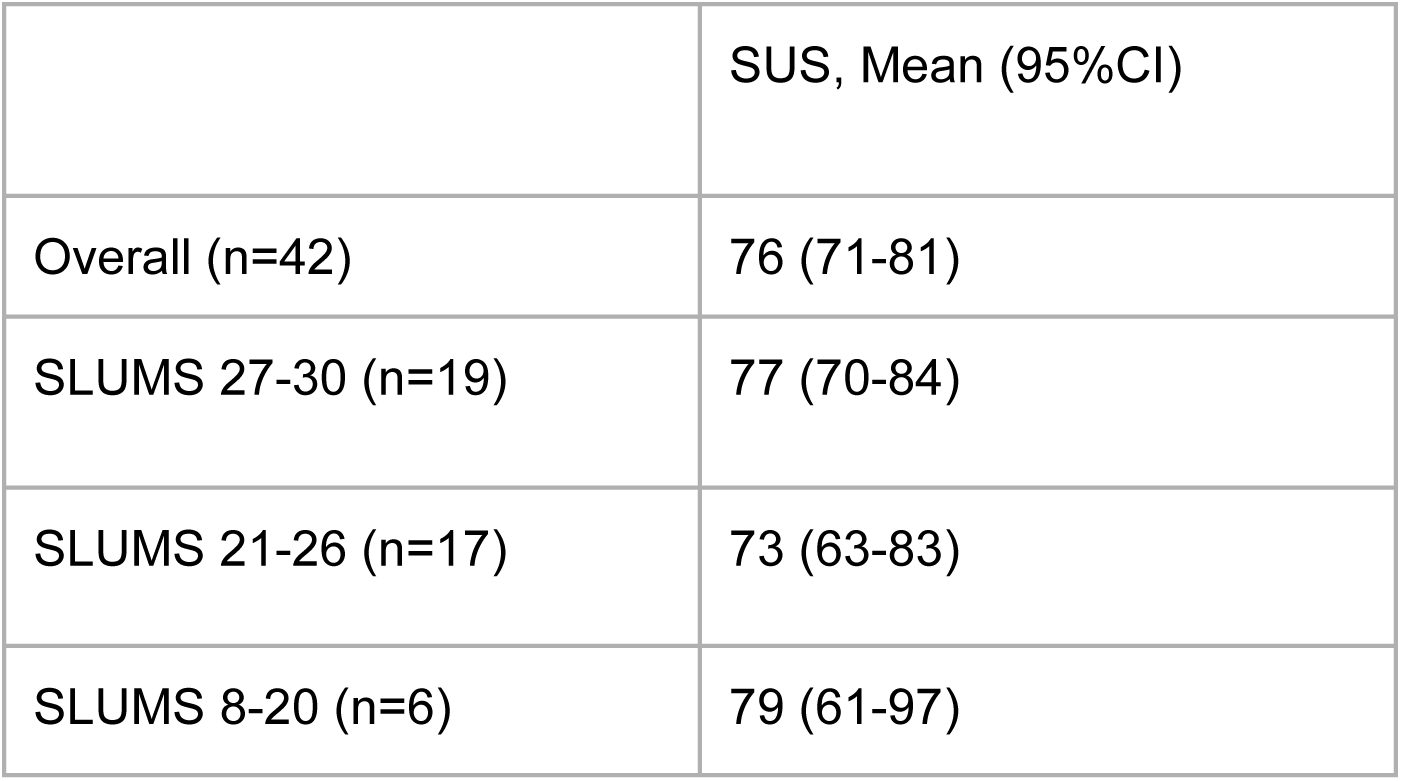
System Usability Scale (SUS) rating by St. Louis University Mental Status Examination (SLUMS) score category.

In the exploratory analysis of the relationship between study completion and SLUMS score category, the Pearson chi-square test was 6.10 with a *P*-value of .047. The median (IQR) SLUMS score was 26 (23–28) in participants who completed the study and 20 (13–24) in participants who withdrew before study completion. In the Receiver Operating Characteristic (ROC) analysis of the SLUMS score as a predictor of study completion, the ROC area under the curve (AUC) was 0.78, with a 95% confidence interval from 0.58-0.97 (Figure 1). A SLUMS score cutpoint of ≥15 correctly predicted study completion in the most (88%) participants, with a sensitivity of 98% and specificity of 38%.

**Figure 1:**
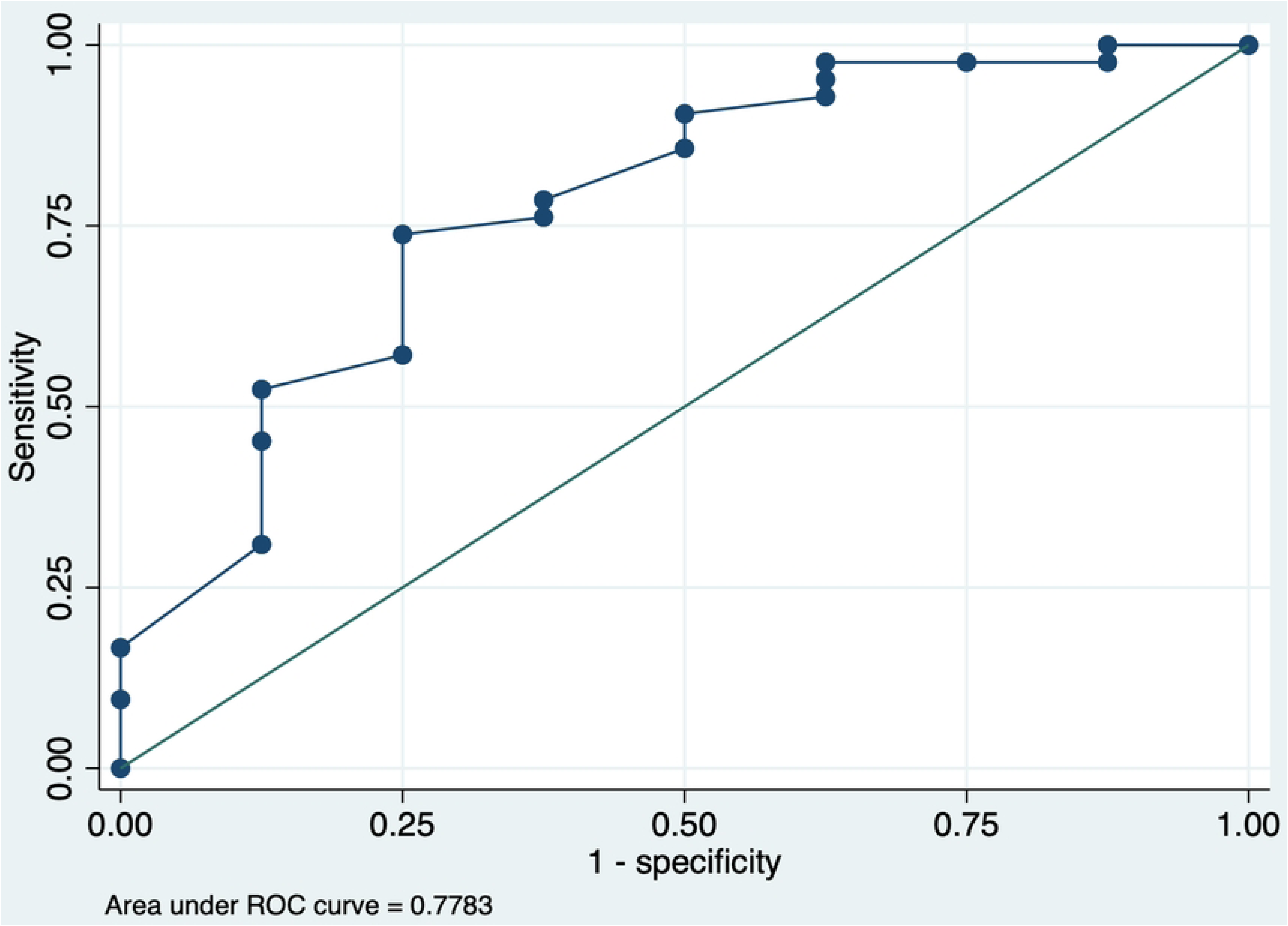
Receiver Operating Characteristic (ROC) curve of St. Louis University Mental Status Examination (SLUMS) score as a predictor of withdrawal from the study before completion. The area under the ROC curve was 0.78.

## DISCUSSION

Participants who completed the study protocol found the prototype to have good usability overall and within categories of SLUMS score. Usability, as represented by the mean SUS rating, was similar across categories of SLUMS score. SLUMS score predicted study completion better than chance, but the ROC AUC was below the conventional lower limit of 0.8 for the ‘moderate’ range.(15) Our study demonstrates that a mobile tablet-based take-home cognitive test could be used by patients in a geriatrics primary care practice across a range of cognitive performance from intact to impaired. We also found that participants with lower SLUMS scores tended to discontinue participation early, suggesting that participants with moderate to severe cognitive impairment did not find the testing platform usable.

In the context of prior research demonstrating that mobile device-based cognitive testing can have acceptable diagnostic performance, our study establishes that a geriatric primary care population considers this testing modality usable. Furthermore, our study demonstrates the usability of cognitive testing on a take-home device in a geriatric primary care setting. Cognitive testing using take-home mobile devices offers an appealing alternative to in-office pen-and-paper cognitive screenings, which strain limited provider time in primary care settings.(2,16) Take-home mobile device-based testing will enable longitudinal assessments, which could address limitations in the diagnostic specificity of tests such as the Montreal Cognitive Assessment.(17) Take-home mobile device-based testing will also enable the incorporation of digital biomarkers such as speech and eye movement parameters into testing protocols, whereby mobile device-based testing could eventually surpass the diagnostic performance of traditional testing methods in diagnosing Alzheimer’s Disease and related dementias.(18)

This work helps establish a broad future role for mobile device-based cognitive testing. Users could interact with such testing platforms in traditional healthcare settings for a one-time screening or infrequent monitoring and at home for screening and more frequent monitoring than what is possible with traditional pen and paper-based tests.

Beyond easing the time/burden of administering every test and making these tests more enjoyable and gamified for patients, the future development of mobile device-based cognitive testing could exceed the sensitivity and specificity of pen and paper tests can provide in that they will measure: 1) Attention (e.g., auditory, sustained vigilance, working memory); 2) Processing speed; 3) Language (e.g., generativity, fluency, object naming); 4) Learning and memory (e.g., free recall and recognition); 5) Executive functioning (e.g., mental flexibility, set shifting, problem-solving, abstract reasoning); and 6) Visual-perceptual reasoning. Mobile device-based cognitive testing could also offer clinicians highly interpretable computer-generated diagnostic reports and a testing experience far more pleasant and game-like than conventional pen and paper cognitive screening tools.

### Limitations

This study tested the usability of a prototype; we have yet to establish the psychometric validity of this particular set of test modules. Several features of our study may limit the generalizability of our findings in important ways. The present study acknowledges a limitation in the sample demographics, characterized by a uniform representation of individuals who self-identified as White and non-Hispanic. This homogeneity in racial and ethnic backgrounds may limit the findings’ generalizability to a broader, more diverse population of older adults. The study’s outcomes and conclusions may not fully account for the variations in cognitive experiences and preferences among individuals from different racial, ethnic, and cultural backgrounds. This study required usable home Wi-Fi service and motor and sensory ability to use the study device, so the results may not be generalizable to older persons without high-speed internet access, or economically disadvantaged persons. We required a legally authorized representative for subjects with questionable capacity for informed consent, so our results may not be generalizable to unbefriended older adults with mild-to-moderate cognitive impairment. Finally, self-selection likely occurred at multiple stages of our recruitment, and our sample should be assumed to represent older adults who are comfortable volunteering for research on cognitive testing and concerned enough to do so, a possible source of healthy user bias. A more diverse participant pool could offer valuable insights into the usability and acceptance of the tablet-based cognitive test across a broader spectrum of older adults. While the findings shed light on the feasibility of this specific demographic, future studies should include a more heterogeneous sample to ensure the robustness and applicability of the results across various populations.

### Conclusion

Take-home mobile device-based cognitive testing is a usable strategy in older adult primary care patients across a range of cognitive function. However, more severe cognitive impairment predicts unwillingness to engage with this technology. Future studies must systematically enroll economically disadvantaged persons, non-English speaking persons, and persons from racial minorities to ensure that results generalize to all potential users so the technology can achieve optimal public health impact. Take-home mobile device-based testing could be part of a flexible cognitive testing and follow-up strategy that includes mobile device-based testing in healthcare settings and pen-and-paper cognitive testing, depending on individual patient characteristics. Mobile device-based cognitive testing has the potential to increase the flexibility and reach of cognitive screening and follow-up for older adults at risk of or diagnosed with Major Neurocognitive Disorder.

## MATERIALS and METHODS

### Study Design and Participants

The purpose of LifeBio Brain Phase 1 was to prospectively assess the usability of a prototype mobile cognitive testing application. We recruited volunteers from an academically-affiliated Geriatric Medicine practice that focuses on primary care of older adults. Advarra provided Institutional Review Board (IRB) approval and oversight of all study materials and procedures through a reliance agreement with the Rhode Island Hospital IRB.

### Ethics Statement

The Advarra Institutional Review Board (IRB) reviewed and approved the study via single-IRB reliance by the Rhode Island Hospital IRB. Participants provided written informed consent, except in cases where chart review or the referring provider indicated cognitive impairment, in which cases a legally authorized representative provided written informed consent and the participant signed written assent.

### Inclusion and Exclusion Criteria

This study included patients of the Geriatric Medicine practice whose most recent SLUMS, Montreal Cognitive Assessment, or Mini Mental Status Examination Score was greater than 15, whose motor, hearing, and vision abilities enabled using a mobile tablet device, and who had a reliable home Wi-Fi service. We recruited patients from November 2, 2022 to February 13, 2023. The principal investigator screened each potentially eligible subject for impaired capacity for informed consent via chart review and direct communication with the referring healthcare providers. We required informed consent from a legally authorized representative for all subjects with impaired capacity for informed consent. We also required signed confirmation of assent from subjects with impaired capacity for informed consent. Subjects were advised of their right to disenroll from the study at any time, for any reason, without any repercussions to their current or future medical care, and with pro-ration of the study financial incentive if they completed some but not all of the protocol. Subjects who completed the protocol received a financial incentive for participation at the end of the protocol.

### Procedures

The principal investigator pre-screened all participants for study eligibility via medical record review under an IRB-approved HIPAA waiver. The principal investigator notified healthcare providers (physicians and nurse practitioners) of all potentially eligible patients scheduled for visits. Referring healthcare providers had knowledge of the study objectives, inclusion, and exclusion criteria, but did not formally screen patients for eligibility.

Referred potential subjects and, when applicable, legally authorized representatives, met in person with a study team member for a concise overview of the study. The overview included a brief introduction to the mobile tablet device and instructions for opening the cognitive testing application. The study team member formally screened the potential subjects for eligibility and then obtained informed consent from the subject or legally authorized representative and, when applicable, signed assent from the subject. The study team then collected demographic information and administered the SLUMS. The study team then provided the study device, including a stylus and a detailed tutorial on using the device, connecting to Wi-Fi, and using the study application.

Participants were instructed to engage with the mobile cognitive testing application twice daily for 5 days - once during the morning and once in the evening. After this period, a member of the study team collected the iPad and administered the SUS.

### Mobile Application

This study tested the usability of a prototype cognitive testing mobile application. The application displayed 7 distinct modules in random order:

1. In ‘Touch the Dot,’ the mobile device displayed a dot moving between random positions on the screen, and instructed participants to ‘touch the dot.’
2. In ‘What is This?,’ the mobile device displayed a series of images and instructed participants to describe what they see on the screen aloud. The app randomly selected 10 images to display from a set of 60.
3. ‘Connect the Circles,’ displayed an adaptation of Trail Making Test A, in which the application instructs participants to touch circles sequentially in numeric order and responds by displaying a trail of line segments.(19)
4. In “Animal Names,” a verbal fluency test, the mobile device displayed a timer and instructed participants to name as many animals as possible.
5. “Draw a Clock,” a digital adaptation of the Clock Drawing Test,(20), instructed participants to draw an analog clock on the touchscreen using a stylus or finger their finger, include all the numbers, and draw the hands of the clock so that the time reads “10 minutes to 11 o’clock.”
6. In “Remember Number,” the device screen displayed a 4-digit number, and instructed the participant to remember it. Subsequent screens instructed the participant to enter the number on a keypad, and then to enter the number on a keypad in reverse order.
7. In ‘Describe the Picture,’ the mobile device displayed a stylized photograph of a meal and instructed the participant to describe what they saw, and then displayed a landscape photograph and instructed the participant to talk about some places they have traveled.

### Outcomes

The primary outcome was the usability of the mobile cognitive testing application, as measured by the SUS. Participants completed the SUS when returning the mobile device to the study team.(14,21) The SUS measures participants’ subjective experience with a digital system or product using 10 Likert items.(22) The instrument alternates between negatively-framed questions and positively-framed questions. The scoring of the SUS is on a scale of 0 to 100.(21) We derived SUS scores from participant responses using methods previously described: we subtracted 1 from the Likert Scale value for questions 1,3,5,7, and 9 and subtracted the Likert Scale value from 5 for questions 2,4,6,8, and 10. We multiplied the resulting sum by 2.5 to obtain the total SUS score.(21) Based on prior studies of SUS interpretation, a score of 65 to 75 indicates ‘good’ usability, and scores above 85 indicate ‘excellent’ usability.(14)

### Statistical Analysis

The primary outcome, the mean SUS score with 95% confidence intervals, was computed using standard methods for normally distributed data. The ROC analysis used nonparametric methods and estimated standard error using the method reported by deLong.(23) All statistical analyses were performed using Stata SE 17.0 (StataCorp, College Station, Texas, USA).

## Data Availability

The authors will consider requests for data sharing

## ACKNOWLEDGEMENTS

The authors would like to acknowledge the individuals who graciously volunteered as subjects in LifeBio Brain Phase 1, and the Brown Medicine Geriatrics Practice whose cooperation made this work possible.

## REFERENCES

1. Chan JYC, Yau STY, Kwok TCY, Tsoi KKF. Diagnostic performance of digital cognitive tests for the identification of MCI and dementia: A systematic review. Ageing Res Rev. 2021 Dec;72:101506.

2. Yokomizo JE, Simon SS, De Campos Bottino CM. Cognitive screening for dementia in primary care: a systematic review. Int Psychogeriatr. 2014 Nov;26(11):1783–804.

3. Manly JJ. Advantages and Disadvantages of Separate Norms for African Americans. Clin Neuropsychol. 2005 May;19(2):270–5.

4. Tolea MI, Chrisphonte S, Galvin JE. The Effect of Sociodemographics, Physical Function, and Mood on Dementia Screening in a Multicultural Cohort. Clin Interv Aging. 2020 Dec;Volume 15:2249–63.

5. Rossetti HC, Lacritz LH, Hynan LS, Cullum CM, Van Wright A, Weiner MF. Montreal Cognitive Assessment Performance among Community-Dwelling African Americans. Arch Clin Neuropsychol. 2016 Oct 25;acn;acw095v1.

6. Rossetti HC, Smith EE, Hynan LS, Lacritz LH, Cullum CM, Van Wright A, et al. Detection of Mild Cognitive Impairment Among Community-Dwelling African Americans Using the Montreal Cognitive Assessment. Arch Clin Neuropsychol. 2019 Aug 28;34(6):809–13.

7. Devlin KN, Brennan L, Saad L, Giovannetti T, Hamilton RH, Wolk DA, et al. Diagnosing Mild Cognitive Impairment Among Racially Diverse Older Adults: Comparison of Consensus, Actuarial, and Statistical Methods. Okonkwo O, editor. J Alzheimers Dis. 2022 Jan 18;85(2):627–44.

8. Zahodne LB, Sharifian N, Kraal AZ, Zaheed AB, Sol K, Morris EP, et al. Socioeconomic and psychosocial mechanisms underlying racial/ethnic disparities in cognition among older adults. Neuropsychology. 2021 Mar;35(3):265–75.

9. Bernstein A, Rogers KM, Possin KL, Steele NZR, Ritchie CS, Kramer JH, et al. Dementia assessment and management in primary care settings: a survey of current provider practices in the United States. BMC Health Serv Res. 2019 Nov 29;19(1):919.

10. Koo BM, Vizer LM. Mobile Technology for Cognitive Assessment of Older Adults: A Scoping Review. Innov Aging [Internet]. 2019 Jan 1 [cited 2023 Aug 3];3(1). Available from: https://academic.oup.com/innovateage/article/doi/10.1093/geroni/igy038/5266911

11. Nef T, Chesham A, Schütz N, Botros AA, Vanbellingen T, Burgunder JM, et al. Development and Evaluation of Maze-Like Puzzle Games to Assess Cognitive and Motor Function in Aging and Neurodegenerative Diseases. Front Aging Neurosci. 2020 Apr 21;12:87.

12. Berron D, Ziegler G, Vieweg P, Billette O, Güsten J, Grande X, et al. Feasibility of Digital Memory Assessments in an Unsupervised and Remote Study Setting. Front Digit Health. 2022 May 26;4:892997.

13. Tariq SH, Tumosa N, Chibnall JT, Perry MH, Morley JE. Comparison of the Saint Louis University mental status examination and the mini-mental state examination for detecting dementia and mild neurocognitive disorder--a pilot study. Am J Geriatr Psychiatry Off J Am Assoc Geriatr Psychiatry. 2006 Nov;14(11):900–10.

14. Bangor A, Kortum PT, Miller JT. An Empirical Evaluation of the System Usability Scale. Int J Hum-Comput Interact. 2008 Jul 29;24(6):574–94.

15. Lasko TA, Bhagwat JG, Zou KH, Ohno-Machado L. The use of receiver operating characteristic curves in biomedical informatics. J Biomed Inform. 2005 Oct;38(5):404–15.

16. Athilingam P, Visovsky C, Elliott AF, Rogal PJ. Cognitive Screening in Persons With Chronic Diseases in Primary Care: Challenges and Recommendations for Practice. Am J Alzheimers Dis Dementiasr. 2015 Sep;30(6):547–58.

17. Davis DH, Creavin ST, Yip JL, Noel-Storr AH, Brayne C, Cullum S. Montreal Cognitive Assessment for the detection of dementia. Cochrane Dementia and Cognitive Improvement Group, editor. Cochrane Database Syst Rev [Internet]. 2021 Jul 13 [cited 2023 Aug 14];2021(7). Available from: http://doi.wiley.com/10.1002/14651858.CD010775.pub3

18. Kourtis LC, Regele OB, Wright JM, Jones GB. Digital biomarkers for Alzheimer’s disease: the mobile/wearable devices opportunity. Npj Digit Med. 2019 Feb 21;2(1):9.

19. Llinàs-Reglà J, Vilalta-Franch J, López-Pousa S, Calvó-Perxas L, Torrents Rodas D, Garre-Olmo J. The Trail Making Test. Assessment. 2017 Mar;24(2):183– 96.

20. Gromisch ES, Beauvais J, Iannone LP, Marottoli RA. Optimizing Clock Drawing Scoring Criteria: Development of the West Haven-Yale Clock Drawing Test. J Am Geriatr Soc. 2019 Oct;67(10):2129–33.

21. Jordan PW, Thomas B, McClelland IL, Weerdmeester B, editors. SUS: A “quick and dirty” usability scale. In: Usability Evaluation In Industry [Internet]. 0 ed. CRC Press; 1996 [cited 2023 Aug 30]. Available from: https://www.taylorfrancis.com/books/9781498710411

22. Peres SC, Pham T, Phillips R. Validation of the System Usability Scale (SUS): SUS in the Wild. Proc Hum Factors Ergon Soc Annu Meet. 2013 Sep;57(1):192–6.

23. DeLong ER, DeLong DM, Clarke-Pearson DL. Comparing the Areas under Two or More Correlated Receiver Operating Characteristic Curves: A Nonparametric Approach. Biometrics. 1988 Sep;44(3):837.

